# Integrating the pattern of negative emotion processing and acute stress response with childhood stress among healthy young adults

**DOI:** 10.1101/2021.05.18.21257390

**Authors:** Jianhui Wu, Yutong Liu, Liang Zhang, Naiyi Wang, Nils Kohn, Hongxia Duan

**Affiliations:** Center for Brain Disorder and Cognitive Science, Shenzhen University, Shenzhen, 518060 China; Shenzhen Institute of Neuroscience, Shenzhen 518057, China; Key Laboratory of Behavioral Science, Institute of Psychology, Chinese Academy of Sciences, Beijing, China; Faculty of Education, Beijing Normal University, Beijing 100875, China; Donders Institute for Brain, Cognition, and Behavior, Radboud University Medical Center, Kapittelweg 29, 6525 EN Nijmegen, The Netherlands

**Keywords:** childhood stress, abuse, neglect, ERPs, Late positive potential, acute stress, heart rate

## Abstract

**Background:** Childhood abuse might impair corticolimbic brain regions, which play a crucial role in emotion processing and the acute stress response. Yet few has explored the relationship between childhood stress, emotion processing and acute stress reactivity despite the overlapping brain regions of the last two. Here, we examine the relationships between distinct dimensions of childhood adversity (i.e., deprivation and threat) and emotion processing as well as acute stress response. Furthermore, we explore whether emotion processing (particularly threatening information), which might be particularly relevant for adaptive stress responding among individuals with experience of childhood abuse, mediates the relationship between childhood stress and acute stress response.

**Method:** Fifty-one young adults completed a free-viewing task to evaluate neural response to negative stimuli measured by late positive potential (LPP) of ERPs (Event-related potentials). In a separate day, heart rate and salivary cortisol were collected during a social-evaluative stress challenge (i.e., TSST, Trier Social Stress Test). After the TSST, childhood trauma questionnaire was measured to indicate the level of abuse (as a proxy of threat) and neglect (as a proxy of deprivation) dimensions.

**Results:** Higher level of childhood abuse (but not neglect) was distinctly related to smaller LPP amplitudes to negative stimuli, as well as smaller heart rate reactivity to acute stress. For these participants, smaller LPP amplitudes linked with smaller heart rate reactivity to acute stress. Furthermore, decreased LPP amplitudes to negative stimuli mediated the relationship between higher level of childhood abuse and blunted heart rate reactivity to stress.

**Conclusions:** Consistent with the dimensional model of childhood stress, our study showed that childhood abuse is distinctly associated with neural as well as physiological response to threat. Furthermore, the blunted neural response to negative stimuli might be the underlying mechanism in which childhood abuse leads to blunted acute stress response. Considering that all the participants are healthy in the present study, the blunted processing of negative stimuli might rather reflect adaptation instead of vulnerability, in order to prevent stress overshooting in the face of frequent early-life threatening experiences.

## Introduction

Exposure to different types of childhood stress, ranging from physical, emotional, sexual abuse, and emotional and physical neglect, is widespread with a prevalence of 30%-53% across the globe (Stoltenborgh et al., 2015). Numerous studies have documented the long-term deleterious effects of childhood stress on psychological and physical health (Clark et al., 2010; Hughes et al., 2017; Rod et al., 2020). However, individuals vary greatly in responses to childhood stress, and many exhibit intact daily functioning and emotional health in adulthood (Teicher et al., 2016). To date, the mechanisms underlying the long-lasting influence of childhood stress on health remain unclear. A putative neurobiological mechanism through which childhood stress may increase the risk for psychopathology is by altering stress response, i.e., the hypothalamic-pituitary-adrenocortical axis (HPA) with end product of cortisol and the autonomic nervous system (ANS) (Danese & McEwen, 2012; Heim et al., 2008; Lupien et al., 2009; Suzuki et al., 2014).

Under the background of childhood stress, recurrent stress would lead to HPA system continuously activated and the endocrine system out of whack (Loannidis et al., 2020). Childhood stress such as social deprivation, trauma, and neglect have been found profound effect on the endocrine system (Carlson & Earls, 1997; De billis et al., 1999; Gunnar & Vazquez, 2001). However, the results summarized in recent meta-analyses are mixed. One meta-analysis reported association between childhood stress and blunted cortisol response (Bunea et al., 2017), and another two meta-analyses failed to detect any significant relationship between these two (Fogelman & Canli, 2018; Lai et al., 2021). Fewer studies have explored the role of ANS, which also responds to stress by exerting more rapid effects through its innervating nerves in many organ systems (Bleker et al., 2018). Mixed results were also found in regard to the ANS acute stress response among childhood adversity (e.g., Heim et al., 2000; Lovallo et al., 2012; Murali & Chen, 2005).

The above evidence suggests that childhood adversity may lead to altered stress response, although the concrete results and the potential mechanisms are still under debate. Emotion processing, which has large overlap brain regions with acute stress reactivity, worth our attention. Previous studies found that childhood stress might disrupt neural networks involved in emotion processing particularly for threating stimuli (Kraaijenvanger et al., 2020; Moreno-López et al., 2020). Consistent with this literature, a lower threshold for threat detection, attention bias toward negative information or increased attention to threat has been found in maltreated children (McCrory et al., 2011; Pollak et al., 2009; Pollak & Sinha, 2002; Pollak & Tolley-Schell, 2003). For example, event-related potential (ERP) studies showed that children with physical maltreatment had larger P3b amplitudes, thus increased attention to angry faces (or voices) compared to normal controls (Pollak et al., 2001; Shackman et al., 2007). Notably, among physically-abused children, increased attention to threat predicted more anxiety symptoms (Shackman et al., 2007), while attention bias away from threat was associated with diagnosis of posttraumatic stress disorder (Pine et al., 2005). Furthermore, the disrupted emotion processing among maltreated children might also persist into adults. Gibb and colleagues (2009) found that young adults with a history of moderate to severe childhood abuse exhibited attention bias as well as interpretation bias to angry expressions. Another study showed that adults with childhood interpersonal trauma exposure failed to differentiate between non-threat and threat-related cues (Chu et al., 2016). Specifically, the amygdala plays a central role in the processing of emotional information including threat detection and appraisal and facilitated attention to salient stimuli (Cunningham & Kirkland, 2014; Murray, 2007; Ohman, 2005; Sergerie et al., 2008; Vuilleumier, 2005). Meanwhile, interacting with the hippocampus and medial prefrontal cortex, the amygdala is a key brain region for stress regulation (Ulrich-Lai & Herman, 2009), as reduced emotion regulation ability revealed as decreased prefrontal cortex activation predicted enhanced responses to acute social stress in cortisol and α-amylase (Kaldewaij et al., 2019).

Altogether, these studies suggest a possible link among childhood stress, emotion processing and acute stress response. However, only a limited number of studies so far have examined these associations. For example, Kircanski et al. (2019) found that in early puberty (mean age 11.37 years), a higher level of childhood stress was related with a decreased cortisol level during a psychosocial stress task, while decreased cortisol was related to increased white matter integrity in the frontolimbic tracts. However, no direct relationship between childhood stress and white matter integrity in the frontolimbic tracts was found. Results from Kaiser and colleagues (2018) showed that among unmedicated young women (mean age 26.41) with variable psychiatric diagnoses, higher severity of threat-related childhood stress was associated with altered resting-state functional connectivity within corticolimbic circuits, which in turn was related to blunted cortisol response to acute stress. However, both these two studies only assessed structure and resting state of brain regions associated with rather than a direct measurement of emotion processing, and they only focused on cortisol response to acute stress. Therefore, in our current study, we used a free viewing paradigm to measure emotion processing with the ERP method and we both measure cortisol (represents HPA axis) and heart rate (represents ANS) in the acute response in healthy young participants. The utilizing of this specific sample was to exclude the possible influence of psychiatric condition and developmental stage (Bunea et al., 2017). For example, adolescents with mild/moderate depression combined with a history of maltreatment showed enlarged cortisol reactivity, but those with moderate/severe depression exhibited blunted cortisol reactivity regardless of childhood maltreatment history (Harkness et al., 2011). The relationship between cortisol reactivity is also found more blunted among adults than children (Bunea et al., 2017).

The late positive potential (LPP), an ERP component that is characterized by a prolonged positive deflection peaking approximately 300-400 ms after the onset of an emotional stimulus is an important marker of emotion processing (Cuthbert et al., 2000). The LPP is sensitive to intrinsically salient stimuli and reflects attention toward affective stimuli (for a review, see Hajcak et al., 2020). Furthermore, source analysis and simultaneous EEG-fMRI recording revealed that neural substrate of the LPP involves cortical and subcortical emotion processing brain regions including the prefrontal cortex and amygdala (De Rover et al., 2012; Keil et al., 2002; Liu et al., 2012; Sabatinelli et al., 2007). Test-retest stability among different population suggests the LPP following emotional stimuli is a stable and reliable marker of emotion processing (Bondy et al., 2018; Kujawa et al., 2013).

According to a recent theoretical model, there are two dimensions underlying diverse childhood stress, i.e., *threat* and *deprivation*, and the two dimensions would have distinct influence on brain and biological systems (McLaughlin et al., 2014). Specifically, threat, referring to interpersonal violence exposures of actual or threatening harm (e.g., physical, emotional and sexual abuse), influences the development of cortico-limbic circuits that underlie fear learning and salience processing; deprivation, referring to the lack of expected cognitive and social inputs as in the case of physical and emotional neglect, influences the development of the association cortex, which in turn might produce difficulties in multiple domains in executive functioning (Sheridan & McLaughlin, 2014; Teicher & Samson, 2016; Teicher et al., 2016). Interestingly, there is some empirical evidence suggesting that childhood experiences of *threat* but not *deprivation* are associated with blunted sympathetic and cortisol responses to acute stress (Busso et al., 2017), as well as with blunted salience processing in terms of self-inflicted error and fear (Wu et al., 2021; Machlin et al., 2019).

To summarize, in the current study, we aimed to investigated whether emotional processing is the underlying mechanism of childhood threat and acute stress response. The link between stress and corticolimbic functioning is bidirectional, such that crucial corticolimbic brain regions (e.g., prefrontal cortex, hippocampus and amygdala) not only are impaired by chronic stress but also play a crucial role in neuroendocrine and autonomic regulation and thus the acute stress response (Herman & Cullinan, 1997; Lupien et al., 2009). Therefore, we predicted that there are significant relationship among childhood threat, emotion processing, and acute stress response, and emotion processing is the underlying mechanism between childhood threat and acute stress response. Specifically, we predicted that neural processing of negative stimuli (indicated by LPP amplitudes) would mediate the effects of childhood threat and acute stress response.

## Method

### Participants

As part of a larger project addressing the predictors of stress reactivity (Chen et al., 2018; Xin et al., 2017, 2020), this study sought to specifically investigate the impact from the perspective of two dimensions of childhood stress (i.e., threat and deprivation) on stress reactivity. Considering potential influences on stress responses, all participants were pre-screened and excluded according to the following criteria: major chronic physiological disease or endocrine disorder; history of psychiatric or neurological disorders; symptoms of chronic anxiety, depression or insomnia; chronic use of psychiatric, neurological, or endocrine medicine; chronic overnight work or irregular day/night patterns; any medication use within three days before participating in the study; current periodontitis; excessive consumption of alcohol (more than two alcoholic drinks a day) or nicotine (more than five cigarettes a day). For females, we included those who did not take oral contraceptives during recruitment and invited them to participate in the experiment before or after their ovulation period (which is defined as the 12th to 16th days prior to the first day of the next menstrual cycle), in order to control the potential influence of sex hormone on stress responses (Kirschbaum et al., 1999; Kudielka & Kirschbaum, 2005). Fifty-two healthy young adults were recruited from the student population of local universities. One male participant was excluded due to the abuse score over 3 standard deviations. Among the remaining 51 participants, there were 19 females and 32 males. The mean age was 22.55 (SD: 1.63), and mean education year was 15.84 (SD: 1.35).

All participants gave written informed consent at the beginning of the experiment and got monetary compensation for their participation. This experiment was approved by the Ethics Committee of Human Experimentation at the Institute of Psychology, Chinese Academy of Sciences.

### General procedure

Participants completed two experimental sessions within two weeks (the interval between the first and second session was within 6 days, except for one participant with a 13-day delay). For the first session, participants completed the passive viewing task with their EEG continuously recorded. For the second session, participants returned to our lab to complete the TSST while their acute stress responses were monitored. The TSST was implemented between 1:00 pm to 5:00 pm to avoid the circadian fluctuation of cortisol levels (Dickerson & Kemeny, 2004; Kudielka et al., 2004) and heart rates (e.g., Vandewalle et al., 2007). Upon arrival, participants were instructed to rest in a quiet room for 30 min during which they filled in demographic information. After the rest period, participants provided the first salivary sample for a baseline measurement. Thereafter, participants completed the TSST task for stress induction, during which heart rate was continuously recorded. Salivary samples were provided at 0 min (post-TSST 1), 20 min (post-TSST 2), 45 min (post-TSST 3), and 60 min (post-TSST 4) after the end of the TSST task. To avoid the negative recalling effect on stress response, we administered the trait anxiety and CTQ questionnaires after the TSST task.

### Passive viewing task

A total of 60 pictures were selected from the International Affective Picture System (IAPS; Lang et al., 1999), of which 30 were negative, depicting unpleasant scenes (e.g., threat and mutilation), and 30 were neutral, depicting neutral scenes (e.g., household objects, leaves, trees). The negative and neutral pictures differed significantly on normative ratings of valence (negative: M = 2.48, SD = 0.57; neutral: M = 5.03, SD = 0.34; t(58) = -21.112, p < .001), arousal (negative: M = 5.66, SD = 0.54; neutral: M = 2.92, SD = 0.49; t(58) = 20.479, p < .001), and dominance (negative: M = 3.77, SD = 0.60; neutral: M = 6.02, SD = 0.36; t(58) = -17.455, p < .001).

After an initial practice block, three experimental blocks were completed with 1-2 min breaks between the blocks. The 60 pictures were presented only once in a random order in each experimental block. Each picture was displayed for 1000 ms in full screen on a 17-in. (43.18-cm) monitor, occupying about 27.3° of horizontal visual angle and about 21.8°of vertical visual angle with a viewing distance of approximately 70 cm. The inter-trial interval varied randomly between 1200 and 1800 ms, during which a white cross was presented in the center of a black background (Hot et al., 2006). Participants were instructed to watch the pictures attentively.

### EEG recording and processing

During the free-viewing task, the EEG was recorded from 64 scalp sites using Ag/AgCl electrodes, which were placed according to the international 10–20 system and mounted in an elastic cap (Neuroscan Inc., Charlotte, North Carolina, USA). The EEG data were processed using Scan 4.3 software (Neuroscan, USA). Eye-movement artifacts were corrected from the EEG data using a regression procedure implemented in the Neuroscan software (Semlitsch et al., 1986). Data were digitally low-pass filtered with 30 Hz and were epoched into periods of 1200 ms (including 200 ms pre-stimulus time as baseline) time-locked to the onset of the emotional pictures. Trials with artifacts exceeding ±100 μV were rejected from analysis.

For each participant, ERP waveforms were averaged for the negative and neutral pictures separately. The LPP was defined as the mean amplitude in the time window of 300-1000 ms at the parietal region (P1, Pz, and P2). The electrodes and time window were chosen according to previous literature (Hajcak & Foti, 2020; Vallet et al., 2020) and our own data which indicated the location of the maximum LPP amplitude.

### Acute stress induction

The TSST task has been shown to be effective in eliciting stress responses (Buchanan et al., 2012, 2009). The task was consisted of a 5-min preparation, a 5-min speech, and a 5-min mental arithmetic. In the preparation period, participants were seated in laboratory room A and instructed to prepare a speech for an imagined scenario in which they were accused of shoplifting and they had to defend themselves in front of the store managers. They were also informed that their performance would be videotaped and evaluated. After preparation, participants were escorted to laboratory room B, where three experimenters (two females) in white coats pretending to be managers were presented. After the speech, participants were asked to complete a mental arithmetic task, which was to subtract serially the number 13 starting at 1022 as fast and accurate as possible. Once an error was made, participants had to restart at 1022. Throughout the speech and arithmetic task, participants spoke into a microphone and a video camera in front of the three experimenters. The experimenters communicated with the participant in a neutral manner with a neutral expression and provided no facial or verbal feedback.

### Measurement of acute stress response

A wireless chest heart rate transmitter and a wrist monitor recorder (Polar RSC800CX, Polar Electro, Finland) was used for heart rate recording. The heart rate was recorded for 5 min at baseline, continuously recorded for 15 min throughout the TSST task, and recorded for 3 min each for the four post-stress measurements at 0 min, 20 min, 45 min, and 60 min after the end of the TSST (i.e., post-TSST 1-4). Averaged heart rate across each recording period was obtained from the Polar performance software and was defined as the number of beats per minutes (bpm). Heart rate values were entered into the analysis as bpm for the different measurement timepoints.

Salivette collection tubes (Sarstedt, Rommelsdorf, Germany) were used to collect saliva samples for salivary cortisol levels at baseline and at 0 min (post-TSST 1), 20 min (post-TSST 2), 45 min (post-TSST 3), and 60 min (post-TSST 4) after the end of the TSST. Saliva samples were frozen at -22 °C immediately after collection until analysis and was thawed and centrifuged at 3000 rpm for 5 minutes before analysis. Cortisol concentration was determined by use of a commercial electrochemiluminescence immunoassay (Cobas e 601, Roche Diagnostics, Numbrecht, Germany) with the lower sensitivity being 0.5 nmol/l. Three cortisol values were missing due to insufficient saliva and imputed by combining the group mean and standard deviation for the missing cortisol sample at that time point, and the mean of the available cortisol samples of the participant (Booij et al., 2013). Cortisol values were entered into the analysis as nmol/l concentrations for the different measurement timepoints.

### Questionnaires

Childhood stress was assessed by a 28-item version CTQ on a 5-point Likert scale (1, never true; 5, very often true) (Bernstein & Fink, 1998). The CTQ is a self-reported questionnaire, which is used as a quantitative measure of the severity of childhood adversity within different populations with or without psychopathology (Viola et al., 2016). The exemplary item is ‘when I was growing up, I got hit so hard by someone in my family that I had to see a doctor or go to the hospital’. Originally, there are five subscales in the CTQ including emotional, physical and sexual abuse, and emotional and physical neglect. According to dimensional model of childhood stress (McLaughlin et al., 2014; Sheridan & McLaughlin, 2014; Teicher & Samson, 2016; Teicher et al., 2016), we calculated abuse total score (as a proxy of threat) by summing up scores from emotional, physical and sexual abuse subscales, and neglect total score (as a proxy of deprivation) by summing up scores from emotional and physical neglect subscales.

Trait anxiety was measured with the Chinese version of the trait subscale of the State-Trait Anxiety Inventory which showed good reliability and validity (STAI-T; Spielberger, 1983; Zhang et al., 2012).

### Data preparation

For the emotion processing, to increase stability of the data, the mean LPP amplitude was calculated by averaging the LPP amplitude across the P1, Pz, and P2 electrodes. To validate the valence effect on the LPP amplitude, a one-way repeated-measures ANOVA with valence (negative vs. neutral) as within-subjects factor was performed. Then, the LPP difference waves (ΔLPP) was calculated by subtracting neutral from negative LPPs to serve as an ERP index of emotional processing.

For the stress reactivity, to evaluate physiological stress reactivity to the TSST, repeated-measures ANOVAs were first performed on salivary cortisol levels and HR with the measurement time point as the within-subject variable. Greenhouse-Geisser correction was employed when sphericity assumption in the ANOVA for repeated measures was violated. Bonferroni correction was used in post-hoc multiple comparisons. Then, heart rate increase (ΔHR) was calculated by subtracting the baseline from the average HR of the TSST. The area under the curve with respect to ground (AUCg) of cortisol was computed to indicate the HPA response to stress (Pruessner et al., 2003).

### Data analysis

Multiple linear regression with bootstrap (n = 1000 times of resampling with replacement) was used to investigate the differential influence of the two dimensions of childhood stress (i.e., threat and deprivation) on emotion processing and acute stress reactivity in our study.

Firstly, for the the relationship between childhood stress and emotion processing *(path 1)*, the primary predictors were abuse and neglect total scores. Sex and trait anxiety were covariate variables. The dependent variables was emotion processing index (i.e., ΔLPP amplitude).

Secondly, for the relationship between childhood stress and acute stress response *(path 2)*, the primary predictors and covariate variables were consistent with the path 1. The dependent variables were acute stress reactivity index (i.e., ΔHR and cortisol AUCg). Furthermore, in order to test the hypothesis that abuse and neglect are distinctly associated with emotion processing and acute stress reactivity, we used a conservative method by examining the overlap of CIs from abuse and neglect. In the event that the CIs overlapped by less than 50% of one CI arm, the beta weights would be considered statistically significantly different from each other (*p* < 0.05, Cumming 2009, 2014). Thirdly, to explore the relationship between emotion processing and acute stress reactivity *(path 3)*, hierarchical regression analyses with bootstrap (n = 1000 times of resampling with replacement) were then conducted to investigate the predicting value of the ΔLPP amplitudes on the stress response (i.e., ΔHR and cortisol AUCg). Sex and trait anxiety were treated as control variables.

Finally, we used the simple mediation model (Model 4) from the PROCESS tool for SPSS (Hayes, 2013) to determine whether emotion processing (ΔLPP amplitude) mediated the association between childhood stress (abuse or neglect) and physiological reactivity (ΔHR or cortisol AUCg) *(path 4)*. Sex and trait anxiety were included as covariates in these models. To determine significance of the indirect effect, PROCESS produces bootstrapped CIs (CIs that do not include zero indicate significant mediation; Hayes 2013). The standard error of the model is estimated by HC3 (Davidson & MacKinnon, 1993).

## Results

### Subjective measurements

The mean score of trait anxiety was 40.20 (SD: 5.41). When the CTQ was subdivided into abuse and neglect, the average abuse score was 16.67 (SD: 1.89) and the average neglect score was 16.78 (SD: 5.51).

### ERP data

The ERP waveforms time-locked to the negative and neutral picture onset and the difference wave (negative minus neutral) are shown in Figure 1. Negative pictures elicited larger LPP amplitudes as compared to neutral pictures (F(1,50) = 112.35, p < .001). The mean amplitude (± SD) of ΔLPP was 2.36 (± 1.59) μV.

**Figure 1:**
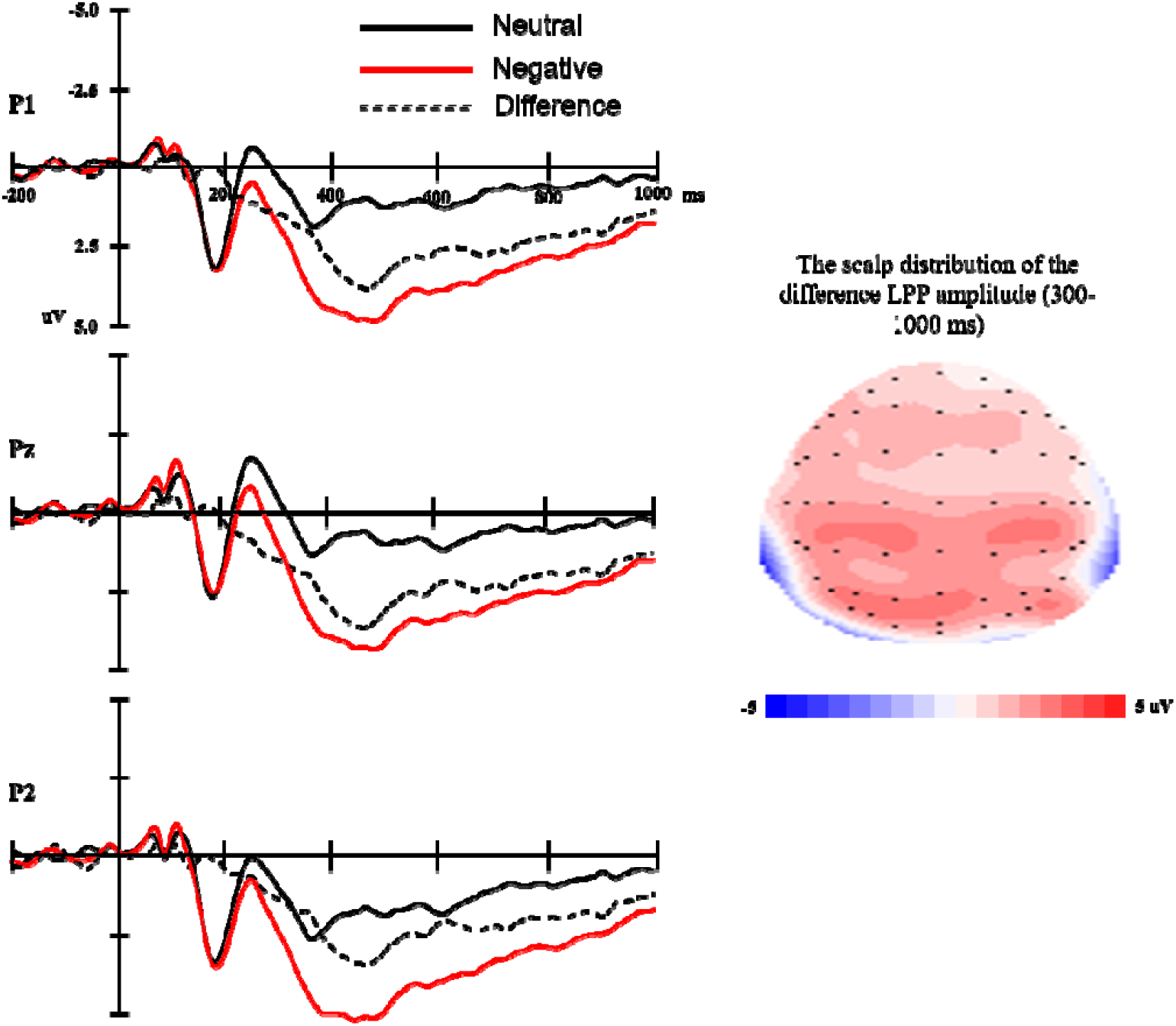
Left: The ERP waveforms time-locked to the negative and neutral picture onset and the difference (negative minus neutral) wave at P1, Pz, P2. Time 0 represents picture onset. Right: The scalp distribution of ΔLPP mean amplitude.

### Stress response

The means and standard deviation of HR and salivary cortisol levels measured before, during, and after the TSST are depicted in Figure 2.

**Figure 2:**
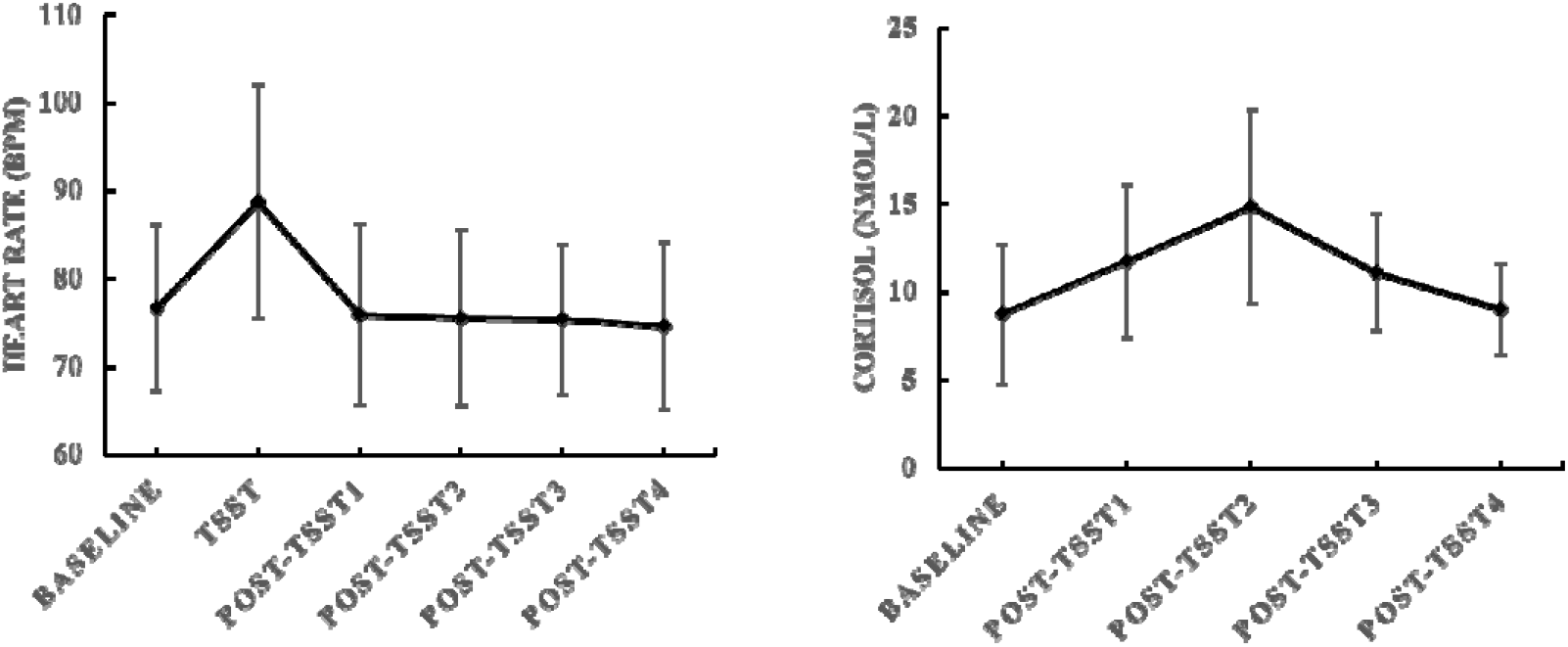
Mean values and standard deviation (SD.) of heart rate (left) and salivary cortisol level (right) measured before, during, and after the TSST task. Baseline: measured after 30-min rest but before TSST; TSST: measured during the TSST task (the mean value of preparation, speech and mental arithmetic); Post-TSST 1 to 4: measured at 0 min, 20 min, 45 min, and 60 min after the end of the TSST task.

For the HR, the repeated-measures ANOVA revealed a significant main effect of Time (F(5, 250) = 63.67, p < 0.01). Post hoc analysis indicated significantly higher HR during the TSST task compared to the baseline and post TSST measures (p_s_ < 0.01). The differences between the baseline HR and the three post-TSST (Post-TSST 1 to 3) measures were not significant (*p_s_* > 0.10). The HR at post-TSST4 was smaller than at baseline (p < 0.05). The mean value (± SD) of the ΔHR was 12.07 (± 8.31).

For the cortisol response, the repeated-measures ANOVA also revealed a significant main effect of Time (F(4, 200) = 40.03, *p* < 0.01). Post hoc analysis showed that salivary cortisol levels measured at 0 min, 20 min, and 45 min after the end of the TSST were significantly higher than cortisol levels measured at baseline (*p*_s_ < 0.01). There was no significant difference between cortisol levels at baseline and that measured at 60 min after the end of the TSST (*p* > 0.10). The cortisol level reached peak at 20 min after the end of the TSST task which was higher than cortisol at 0 min and 45 min after the end of the TSST task (*p*_s_ < 0.01). The mean value (± SD) of AUCg was 19.58 (± 5.74).

### The correlation between two dimensions of childhood stress, emotion processing and acute stress response

#### Path 1: the relationship between childhood stress and emotion processing

For the emotion processing, a multiple linear regression (Table 1) was conducted with predictors of abuse and neglect and control variables (sex, and trait anxiety). The model explained 12% of the variance in ΔLPP amplitudes (R^2^ = 0.12, F(4,50) = 1.57, *p* > 0.10), and only abuse score but not neglect significantly negatively predicted ΔLPP amplitudes (Table 1). To evaluate the hypothesis more precisely, standardized beta coefficients and CIs were compared between CTQ-abuse and CTQ-neglect. As shown in Figure 3 Left, CIs of ΔLPP mean amplitudes for abuse were clearly distinguished from neglect (less than 50%), indicating that the relationships for ΔLPP amplitudes were significantly different between CTQ-abuse and CTQ-neglect.

**Table 1.** Bootstrapping regression of CTQ’s two dimensions on ΔLPP amplitudes.

| Predictors | Standardized regression |  | Standard error (bootstrap) | *p* |
| --- | --- | --- | --- | --- |
|  | coefficient B | 95% CI |  |  |
| Sex | -0.02 | [-0.34, 0.25] | 0.15 | 0.88 |
| Trait anxiety | 0.26 | [-0.07, 0.64] | 0.18 | 0.16 |
| Abuse | -0.51 | [-0.96, -0.15] | 0.21 | 0.02 |
| Neglect | -0.02 | [-0.27, 0.42] | 0.17 | 0.52 |

**Figure 3.**
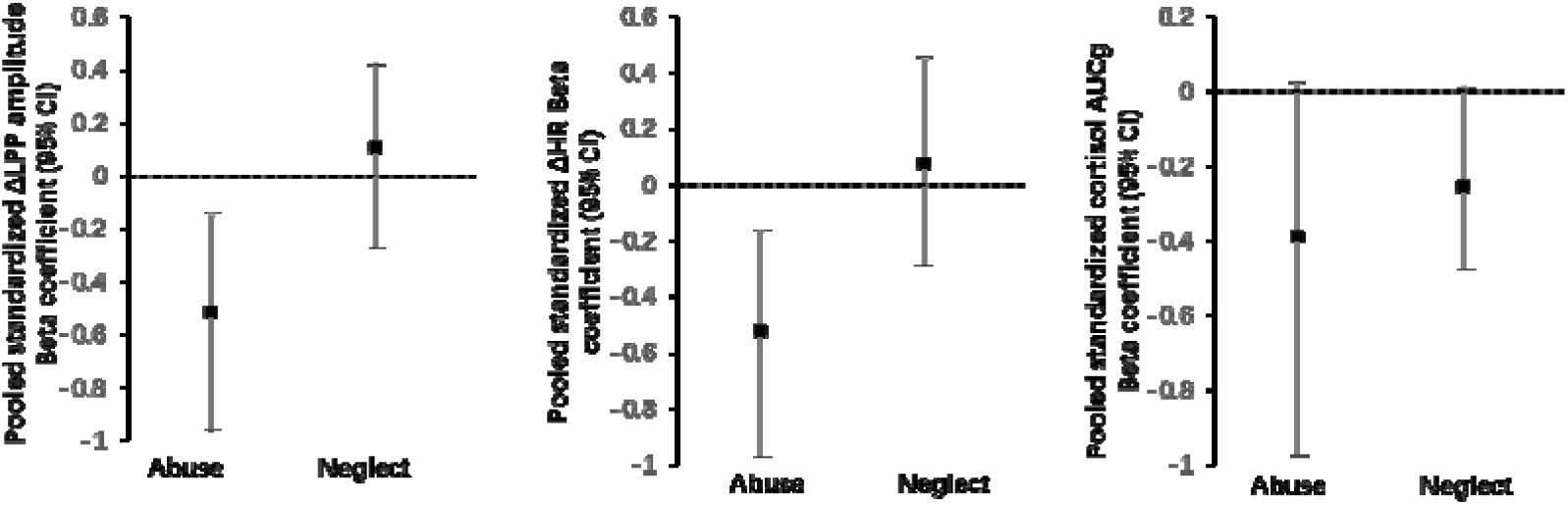
Left: Pooled standardized beta coefficients (with 95% CIs), representing the magnitude of ΔLPP amplitudes, by CTQ-abuse and CTQ-neglect. Middle: Pooled standardized beta coefficients (with 95% CIs), representing ΔHR, by CTQ-abuse and CTQ-neglect. Right: Pooled standardized beta coefficients (with 95% CIs), representing cortisol AUCg, by CTQ-abuse and CTQ-neglect.

#### Path 2: the relationship between childhood stress and acute stress response

For the heart rate reactivity to TSST, the multiple regression model explained 16.5% of the variance of ΔHR (R^2^ = 0.165, F(4,50) = 2.27, *p* = 0.08), and only abuse score but not neglect significantly negatively predicted ΔHR (Table 2). Furthermore, as shown in Figure 3 Right, CIs of ΔHR for abuse were distinguished from neglect (less than 50%), demonstrating that the regression coefficient for CTQ-abuse was considered significantly different from the CTQ-neglect regression coefficient.

**Table 2.** Bootstrapping regression of CTQ’s two dimensions on ΔHR

| Predictors | Standardized regression |  | Standard error (bootstrap) | *p* |
| --- | --- | --- | --- | --- |
|  | coefficient B | 95% CI |  |  |
| Sex | 0.17 | [-0.32, 0.24] | 0.12 | 0.16 |
| Trait anxiety | -0.16 | [-0.09, 0.41] | 0.16 | 0.32 |
| Abuse | -0.52 | [-0.97, -0.17] | 0.21 | 0.02 |
| Neglect | 0.08 | [-0.29, 0.45] | 0.20 | 0.70 |

For the cortisol reactivity to TSST, the regression model was significant and explained 20.9% of the variance of cortisol AUCg (R^2^ = 0.209, F(4,50) = 3.04, p = 0.03). The 95% CIs showed that neglect marginally negatively predicted cortisol reactivity (see Table 3). However, the overlapping CIs analysis showed that CIs of cortisol AUCg for neglect were not significantly distinguished from abuse (more than 50%), indicating that the relationships for cortisol AUCg were not different between CTQ-abuse and CTQ-neglect.

**Table 3.** Bootstrapping regression of CTQ’s two dimensions on cortisol AUCg

| Predictors | Standardized regression |  | Standard error (bootstrap) | *p* |
| --- | --- | --- | --- | --- |
|  | coefficient B | 95% CI |  |  |
| Sex | -0.03 | [-0.29, 0.28] | 0.14 | 0.83 |
| Trait anxiety | -0.06 | [-0.31, 0.27] | 0.14 | 0.62 |
| Abuse | -0.39 | [-0.97, 0.02] | 0.25 | 0.11 |
| Neglect | -0.25 | [-0.47, 0.01] | 0.13 | 0.04 |

#### Path 3: the relationship between emotion processing and acute stress response

Table 4 shows the results of the regression analysis with bootstrap for the HR response to stress with ΔLPP amplitude being predictor and ΔHR being outcome. The model explained 17% of the variance (R^2^ = 0.17, F(3, 50) = 3.20, *p* < 0.05) in the HR response to the acute psychosocial stressor (ΔHR), and ΔLPP amplitude significantly positively predicted ΔHR (see Table 4).

In contrast, the hierarchical regression model for the cortisol response to stress (AUCg) was not significant (R^2^ = 0.03, F(3, 50) = 0.52, *p* > 0.10) and the amplitude of ΔLPP could not predict the AUCg (95% CI: [-0.26, 0.26], *p* > 0.10).

**Table 4.** Hierarchical regression analyses with bootstrap predicting HR response to the stressor.

| Predictors | Standardized regression |  | Standard error |  |
| --- | --- | --- | --- | --- |
|  | coefficient B | 95% CI | (bootstrap) | *p* |
| Sex | -0.12 | [-0.40, 0.14] | 0.14 | 0.38 |
| Trait anxiety | -0.32 | [-0.56, -0.05] | 0.13 | 0.02 |
| ΔLPP amplitude | 0.34 | [0.11, 0.58] | 0.12 | 0.01 |

#### Path 4: Mediation analysis

We first found that only the level of childhood abuse is significantly associated with blunted emotion processing (ΔLPP amplitudes) and blunted heart rate response to stress (ΔHR). Then, we ran a mediation model to test whether emotion process mediated the association of childhood abuse and HR reactivity to the TSST. Adjusting for sex and trait anxiety, a significant indirect effect of abuse through blunted emotion processing to HR reactivity was observed (95%CI: -0.325, -0.002) (Figure 4). After including Δ LPP amplitude as a mediator, the direct effect from abuse to heart rate response became non-significant. Therefore, higher level of childhood abuse predicting smaller heart rate response to acute stress was fully mediated by blunted processing of negative stimuli.

**Figure 4.**
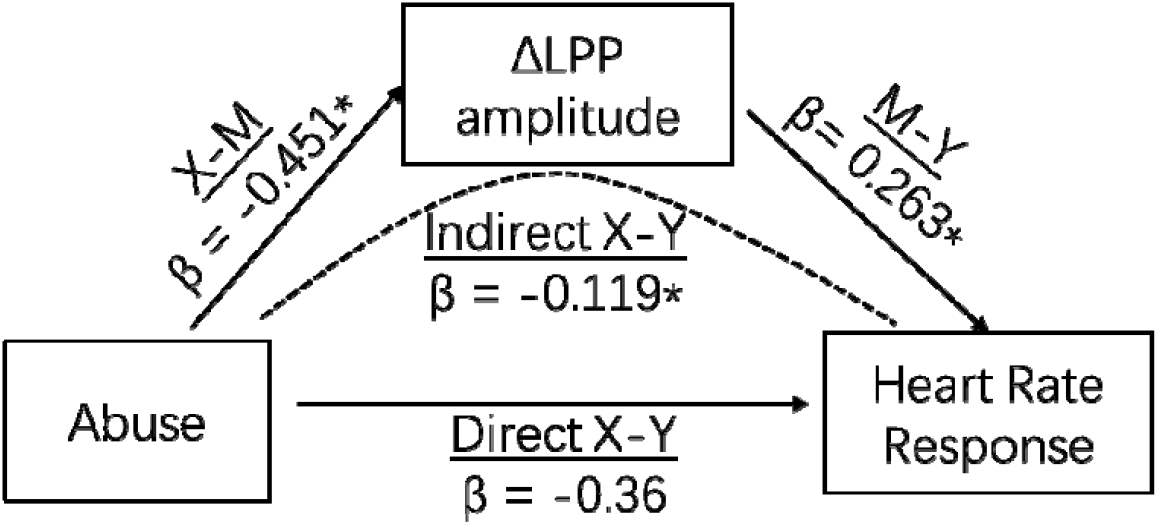
Neural processing of negative stimuli (indicated by ΔLPP amplitudes) mediates the effects of childhood abuse and heart rate response to acute social stress. \**p* ≤ 0.05. LPP: Late positive potential

## Discussion

In this study, we found that higher level of childhood abuse (as a proxy of threat) but not neglect (as a proxy of deprivation) was distinctly associated with blunted neural reactivity to negative stimuli (as indicated by smaller LPP amplitudes) on the one hand, and with blunted HR response to acute psychosocial stress on the other hand. Smaller LPP amplitudes was also related with blunted HR acute stress response. Furthermore, the mediation model showed that childhood abuse predicted blunted HR response through blunted neural reactivity to negative stimuli. These results even hold after controlling sex and trait anxiety, in a sample of healthy young adults.

First of all, we found that enhanced LPP amplitudes to negative compared to neutral pictures in a non-stressful state were associated with higher HR responses to acute psychosocial stress. The LPP is a reliable marker reflecting elaborated attention toward affective stimuli Bondy et al., 2018; Hajcak et al., 2020; Kujawa et al., 2013), which involves cortical and subcortical emotion processing brain regions including the prefrontal cortex and amygdala (De Rover et al., 2012; Keil et al., 2002; Liu et al., 2012; Sabatinelli et al., 2007). The amygdala, as an important part of the limbic system, is well positioned to control basic autonomic arousal processes through the hypothalamus and brainstem circuits (LeDoux 2000), and generates visceral signs of emotional arousal—e.g., changes in heart rate (Critchley et al., 2005; Kuniecki et al., 2002). Prior studies using simultaneous fMRI and EEG recording (Liu et al., 2012; Sabatinelli et al. 2007; 2013), genetic analysis, and pharmacological manipulation (de Rover et al., 2012) have suggested that the increased LPP amplitude reflects amygdala modulation, which plays an important role in autonomic stress response (e.g., Fortaleza et al., 2012). Consistently, we found that enhanced LPP amplitudes to negative compared to neutral pictures were associated with higher HR responses to acute psychosocial stress. It adds to the evidence that negative bias plays a causal role in stress vulnerability (e.g., Fox et al., 2011). For example, prior fMRI and EPR studies found that relatively exaggerated amygdala reactivity and increased LPP amplitude towards negative stimuli before trauma exposure were predictive to posttraumatic symptoms in response to traumatic stressors (Admon et al., 2009; Lobo et al., 2014; Swartz et al., 2015). Egloff and colleagues (2002) found that attentional bias to negative stimuli measured by response times positively predicted HR and blood pressure responses to an evaluated speech task in females. Thus, a generally hypersensitive amygdala may explain both larger LPP amplitudes towards negative stimuli and higher HR response to stress.

When adding the factor of childhood stress, our results showed that childhood abuse is associated with smaller LPP amplitudes to negative stimuli as well as smaller HR reactivity to acute stress. As the theoretical model has proposed, childhood threat as in abusive conditions may differentiate in shaping neurobiological development, such that abuse exposure influences the development of cortico-limbic circuits that underlie salience processing (McLaughlin et al., 2014; Sheridan & McLaughlin, 2014; Teicher & Samson, 2016; Teicher et al., 2016), in turn, modifying physiologic responses to stress (Busso et al., 2017). Consistent with the model, we found that childhood abuse but not neglect was distinctly related with blunted processing to negative stimuli on the one hand, and with blunted heart rate response to acute stress on the other hand. This effect, to some extent, was in line with previous studies that abused children tended to avoid negative information shown as biased attention away from threat (Pine et al., 2005), and as decreased activation of amygdala to negative stimuli (Puetz et al., 2016). Meanwhile, the current result also echoed with one previous study from our own lab that childhood abuse rather than neglect was associated with blunted motivational evaluation of self-inflicted errors among healthy young males (Wu et al., 2021). Furthermore, our mediation analysis showed that childhood abuse predicted blunted HR response through blunted processing to negative stimuli. Considering that the role of amygdala sensitivity in both emotion processing and autonomic stress response as we discussed above, the blunted amygdala with higher level of childhood abuse might be the underlying neural mechanism. Specifically, individuals with higher level of childhood abuse might show diminished amygdala response revealed as smaller LPP reactivity to negative stimuli, which further lead to smaller autonomic response to acute psychosocial stressor-the universal negative event.

Given the healthy characteristic of our participants, results of blunted salience processing in the current study could be explained by the cortical and cognitive adaptation mechanism, i.e., following iterative exposure to threat, the response to negative events might become habituated or downregulated in order to prevent stress overshoot (Heim et al., 2013). Accordingly, a recent scoping review showed that resilient adults after childhood adversity (i.e., those reporting childhood maltreatment without psychopathology) demonstrated improved ability to regulate emotions through medial prefrontal cortex-amygdala downregulation, lower hippocampal activation to emotional faces, and increased amygdala habituation to stress (Moreno-López et al., 2020). Similarly, a study found that higher trait resilience was related with blunted negative emotion processing indicated by smaller LPP amplitudes (Chen et al., 2018). In other words, our study suggested that dampened threat processing (indicated by smaller LPP reactivity to negative stimuli) might rather reflect resilience to childhood abuse in young adults, and this dampened threat processing further leads to diminished stress responses (indicated by smaller HR response to acute stress).

There are some limitations to the current study. First, we only used the retrospective self-reported questionnaire, which might not be a precise way to measure childhood stress. Participants with enhanced emotion processing might have a negative recall bias, in turn, report more severity of childhood stress. By controlling trait anxiety level, we might be able to, to some extent, partial out the effects of personality on recall bias and stress response. Furthermore, although the CTQ is a commonly used instrument for assessment of childhood stress, it does not include the information about the time the adversity occurred. Therefore, it limits our understanding of the specific developmental stage (s) of adversity exposure, which is a crucial factor on the effect of childhood stress on brain and cognitive development (McLaughlin et al., 2014; Teicher et al., 2016). Future studies with retrospective method should consider more sophisticated instrument like the “Maltreatment and Abuse Chronology of Exposure” (Teicher & Parigger, 2015).

In conclusion, building on the previous work, the current study explored the two dimensions of childhood stress on negative emotion processing and acute stress response among the same group of healthy young adults. The results revealed that the severity of childhood abuse is uniquely associated with blunted emotion processing and blunted acute stress response, and blunted emotion processing significantly mediates the relationship between childhood abuse and acute stress response. Considering that all the participants are healthy, the blunted processing to negative stimuli might reflect adaptive responses to prevent stress overshooting in the face of frequent early-life threatening experiences.

## Data Availability

Data is available upon request

